# Admission criteria in critically ill COVID-19 patients: a physiology-based approach

**DOI:** 10.1101/2021.05.30.21257382

**Authors:** Samuele Ceruti, Andrea Glotta, Maira Biggiogero, Pier Andrea Maida, Martino Marzano, Patrizia Urso, Giovanni Bona, Christian Garzoni

## Abstract

**Introduction:** The COVID-19 pandemic required a careful management of intensive care unit (ICU) admissions, to reduce ICU overload while facing resources’ limitations. We implemented standardized, physiology-based, ICU admission criteria and analyzed the mortality rate of patients refused from the ICU.

**Materials and Methods:** COVID-19 patients proposed for ICU admission were consecutively analyzed; Do-not-resuscitate patients were excluded. Patients presenting a SpO_2_ lower than 85% and/or dyspnea and/or mental confusion resulted eligible for ICU admission; patients not presenting these criteria remained in the ward with an intensive monitoring protocol. Primary outcome was both groups’ survival rate. Secondary outcome was a sub analysis correlating SpO_2_ cutoff with ICU admission.

**Results:** From March 2020 to January 2021, 1623 patients were admitted to our Center; 208 DNR patients were excluded; 97 patients underwent intensivist evaluation. The *ICU-admitted* group mortality rate resulted 15.9% at 28 days and 27% at 40 days; the *ICU-refused* group mortality rate resulted 0% at both intervals (p < 0.001). With a SpO_2_ cut-off of 92%, the hypoxia rate distribution did not correlate with ICU admission (p = 0.26); with a SpO_2_ cut-off of 85%, a correlation was found (p = 0.009). A similar correlation was also found with dyspnea (p =0.0002).

**Conclusion:** In COVID-19 patients, standardized ICU admission criteria appeared to reduce safely ICU overload. In the absence of dyspnea and/or confusion, a SpO_2_ cutoff up to 85% for ICU admission was not burdened by negative outcomes. In a pandemic context, the SpO_2_ cutoff of 92%, as a threshold for ICU admission, needs critical re-evaluation.

## INTRODUCTION

In a dramatic situation such as the COVID-19 pandemic, a careful definition of the intensive care unit (ICU) admission criteria is required (1), with the aim to avoid inappropriate resources abuse and to provide adequate patient-tailored management. Clinicians should in fact distinguish between patients who will benefit from ICU admission and those who are unlikely to benefit from it, in order to avoid inappropriately invasive and traumatic measures in those at high risk for poor outcome despite intensive treatment (1, 3, 4). In this regard, triage regulations based on appropriate and accepted ethical principles have been developed in Switzerland, in order to reserve ICU admission only to those who will actually benefit from an intensive medical intervention (2).

Due to the out-of-ordinary pandemic situation, with the concomitant lack of human and material resources, it was relevant to define clear guidelines for ICU admission, with rules respecting ethical principles and structured on the health-system specific medical resources and ICU limits (4). As a consequence of this peculiar setting, restricting decisions were necessary (5).

We conceived and implemented a standardized procedure in our COVID-19 Center in South Switzerland, based on well-determined criteria for ICU admission, which were mainly based on patient’s respiratory pathophysiology. Aim of this study was to analyze the mortality rate and the clinical characteristics of patients assessed for eventual ICU admission, based on these established criteria.

## MATHERIALS AND METHODS

### Study population and data

A retrospective analysis was conducted on consecutive patients with acute respiratory distress syndrome due to COVID-19 pneumonia, triaged for ICU admission from March 2020 to January 2021. According to WHO guidelines (6), SARS-COV-2 laboratory confirmation was defined as a positive result of real-time reverse transcriptase-polymerase chain reaction (RT-PCR) on nasal and pharyngeal swabs. After in-hospital admission, according to the Swiss Academy of Medical Sciences (SAMS) criteria (5, 7), patients were assigned a Do Not Resuscitate (DNR) order if they satisfied the following criteria: endotracheal intubation refusal, hypoxia-related cardiocirculatory arrest, ongoing metastatic oncological disease, end-stage neurodegenerative disease and severe, irreversible chronic diseases like heart failure NYHA IV, COPD GOLD D, liver cirrhosis Child-Pugh > 8, and severe dementia. These patients were consequently excluded from the possibility of ICU admission and were instead followed by specialists in palliative care, as well as treated according to current standards (8). They were not further referred for consultation regarding ICU admission during their hospital stay.

For a standardized evaluation of patients admitted to the hospital who resulted eligible for ICU admission, the Early Warning Score (EWS) was applied by nursing and medical staff (8, 9). The daily frequency of EWS evaluation was performed based on the patients’ clinical condition: for EWS less than 4, the evaluation was performed four times a day, while for EWS greater than 5, the evaluation was performed up to twice an hour (8). For EWS equal or greater than 7, an Intensivist consultation was required.

### ICU evaluation criteria

With the aim to quickly identify patients with worsened clinical conditions (5, 10) and to avoid ICU overload, the Intensivist Consultant evaluated patient’s symptoms, peripheral oxygen saturation (SpO_2_), blood gas analysis values and clinical status. Patients with a partial respiratory failure with a SpO_2_ lower than 85% and dyspnea (or mental confusion) and/or patients with dyspnea or mental confusion alone were eligible for ICU admission (*ICU-admitted* group). Patients admitted to the ICU underwent oro-tracheal intubation and mechanical ventilation (MV) as standard of care. Patients who did not meet the previously mentioned ICU inclusion criteria (*ICU-refused* group) were followed in their clinical course until the ICU criteria were met or a clinical improvement was achieved. DNR patients, as previously mentioned, were not included in these evaluations. The mortality rate of both groups at 28 and 40 days was compared thereafter. Demographics, clinical data and laboratory/radiological results collected during patient’s hospitalization were extrapolated from electronic health records.

With the aim to determine whether the *ICU-admitted* group was a representative sample of the entire ICU patient group, a comparison between the *ICU-admitted* group (patients admitted in ICU exclusively from the intensivist consultation) and the *whole-ICU* population (all ICU patients admitted from intensivist consultation, from other hospitals and from the emergency department, ED) were further performed.

### Outcomes

Primary endpoint was the determination of the survival rate in *ICU-refused* and *ICU-admitted* groups at 28 and 40 days, further comparing the mortality rate between two groups. Secondary endpoint was a comparison between the two groups in relation to clinical and biological aspects, in order to determine any predictive variables associated with ICU admission. These included demographic characteristics (such as age, gender, body-mass-index - BMI), comorbidities (such as arterial hypertension, ischemic heart disease, diabetes, obstructive sleep apnea syndrome – OSAS - and chronic obstructive pulmonary disease - COPD), hemodynamic and respiratory parameters (systolic and diastolic blood pressure, heart rate, temperature, lactate, SpO_2_, partial pressure of oxygen - pO_2_, partial pressure of carbon dioxide -pCO_2_, the need for oxygen therapy and the presence of dyspnea at ICU admission). A specific analysis on patients’ hypoxia distribution between two groups according to SpO_2_ cut-offs of 92% vs 85% was also performed.

### Statistical analysis

Descriptive statistic was performed to summarize the collected clinical data. Data were presented as mean (SD) or median (IQR) for continuous variables, according to data distribution, and as absolute number (and percentage) for categorical variables; data distribution was verified by Kolmogorov-Smirnov test and Shapiro–Wilk test. Differences between patient outcomes were studied by t-test for independent groups or by Mann-Whitney test if non-parametric analysis was required. Similarly, comparison of clinical evolution over time was performed by t-paired test or by non-parametric Wilcoxon test, depending on data distribution. Study of differences between groups of categorical data was carried out by Chi-square statistics. In order to calculate a posterior probability to ICU admission according to clinical binomial data used for patients selected during ICU consultation, a Bayesian analysis of contingency tables were performed. Kaplan-Meier analysis was used to study patients’ survival with the Cox-Mantel log-rank test to ascertain differences between the groups, analyzing all event by time to ICU admission. All Intervals of Confidence (CI) were established at 99%; significance level was established to be < 0.01. Statistical data analysis was performed using the SPSS.26 package (SPSS Inc., Armonk, NY; USA).

### Ethics Committee permissions

This study has been approved by the Ethics Committees of Canton Ticino (Comitato Etico Cantonale, CE_TI_3807), according to the local Federal rules. No funding has been required.

## RESULTS

During the study period 1623 patients were admitted to our COVID-19 center (Figure 1); two-hundred and eight DNR patients were excluded from the analysis. Of the remaining 1415 patients, 100 (7%) underwent Intensivist Consultation during their hospitalization; three (3%) patients refused to give informed consent to their data treatment and were therefore excluded (Figure 1). At consultation, mean patients’ age was 69 ± 11 years (31 – 93), with a median SpO_2_ of 89% (85 – 92), a median pO_2_ of 59.1 mmHg (47.8 – 73.7), a median pCO_2_ of 34.7 mmHg (32 – 39.2) and a median hemoglobin of 13.8 g/L (11.9 – 15.1). Of the 100 patients undergoing Intensivist consultation, sixty-three (64.9%) presented one or more ICU admission criteria and were therefore admitted to the ICU (*ICU-admitted* group), while thirty-four (35%), who did not present the aforementioned criteria (*ICU-refused* group), remained under strict follow-up out of ICU (Figure 1). Demographic and clinical data of both groups are summarized in Table 1.

**FIGURE 1.**
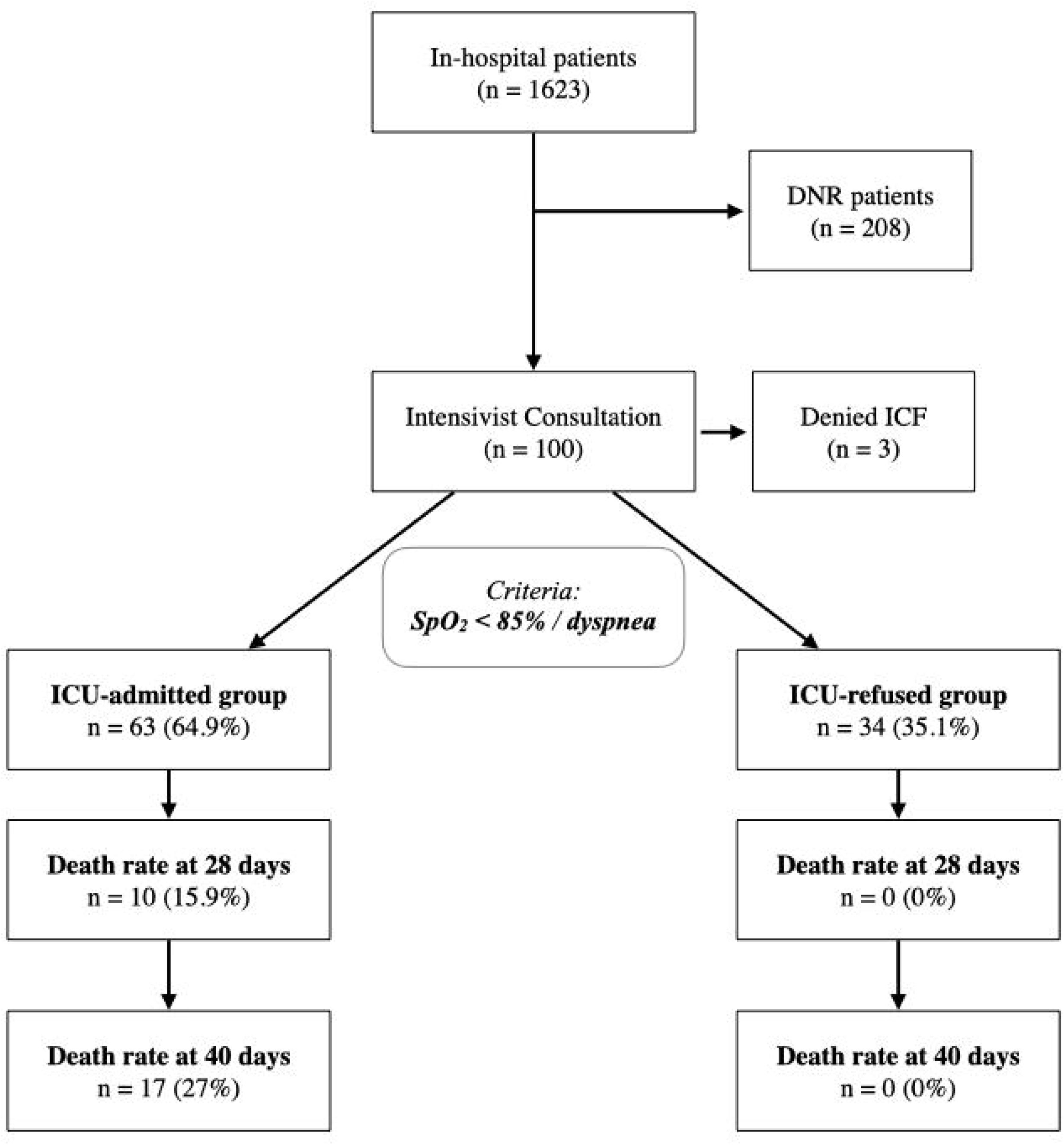
Management of COVID-19 patients evaluated at our COVID-19 center from March 2020 to January 2021. ICU admission was allowed according to standardized criteria. Patients not admitted followed according to EWS score, to quickly identify any change in their medical status. Death rate at 28 and 40 days is reported.

In ICU, 15 additional patients were directly transferred from other hospitals or emergency department (ED) already on MV; the group of the *whole-ICU* patients resulted composed by 81 patients that resulted equivalent comparing to the *ICU-admitted* group (Table SM1).

### Clinical and biological comparison

In the *ICU-admitted* versus *ICU-refused* comparison, no significant difference was found in relation to age (p = 0.87), BMI (p = 0.63), pO_2_ (p = 0.07), pCO_2_ (p = 0.81), lymphocytes count (p = 0.19), CRP level (p = 0.82), creatinine kinase level (p = 0.26), total bilirubin (p = 0.21), lactate (p = 0.20) and hemoglobin (p = 0.69, Table 1). Patients refused from ICU admission resulted more prone to be female (47% vs 19.7%, p < 0.001), with lower LDH (median 375 U/L vs 598 U/L, p = 0.05) and higher platelets count (median 264 vs 198 G/L, p = 0.03). No significant difference was found concerning the presence of ischemic cardiomyopathy (p = 0.57), diabetes (p = 0.69), and COPD (p = 0.59), nor concerning hemodynamics parameters at admission, such as systolic and diastolic blood pressure (p = 0.61 and 0.58 respectively) and body temperature (p = 0.15). All data were reported in Table 1. Patients admitted to the ICU were instead more prone to be affected by arterial hypertension (66.7% vs 47.1%, Chi-square 10.7405, df = 1, p < 0.001).

### Survival rate analysis

At 28 days, the *ICU-refused* group mortality rate was 0%; all *ICU-refused* patients’ clinical condition eventually improved and at forty days from the intensivist consultation some of the patients were already discharged from the hospital. For *ICU-admitted* group, the 28-days mortality rate was 15.9% (10 patients), increasing up to 27% (17 patients) at 40 days (Figure 1). Comparing the 28-days survival rate, a significant imbalance in the distribution of mortality was found in favor of the *ICU-refused* group (Chi-square of 7.195, df = 1, p = 0.007, Figure 2A); extending the comparison at 40 days, a further imbalance was found, with a 25.7% (17 patients) and 0% (no patients) of mortality rate respectively in *ICU-admitted* and *ICU-refused* groups (Chi-square 13.2136, df = 1, p < 0.001, Figure 2B).

**FIGURE 2.**
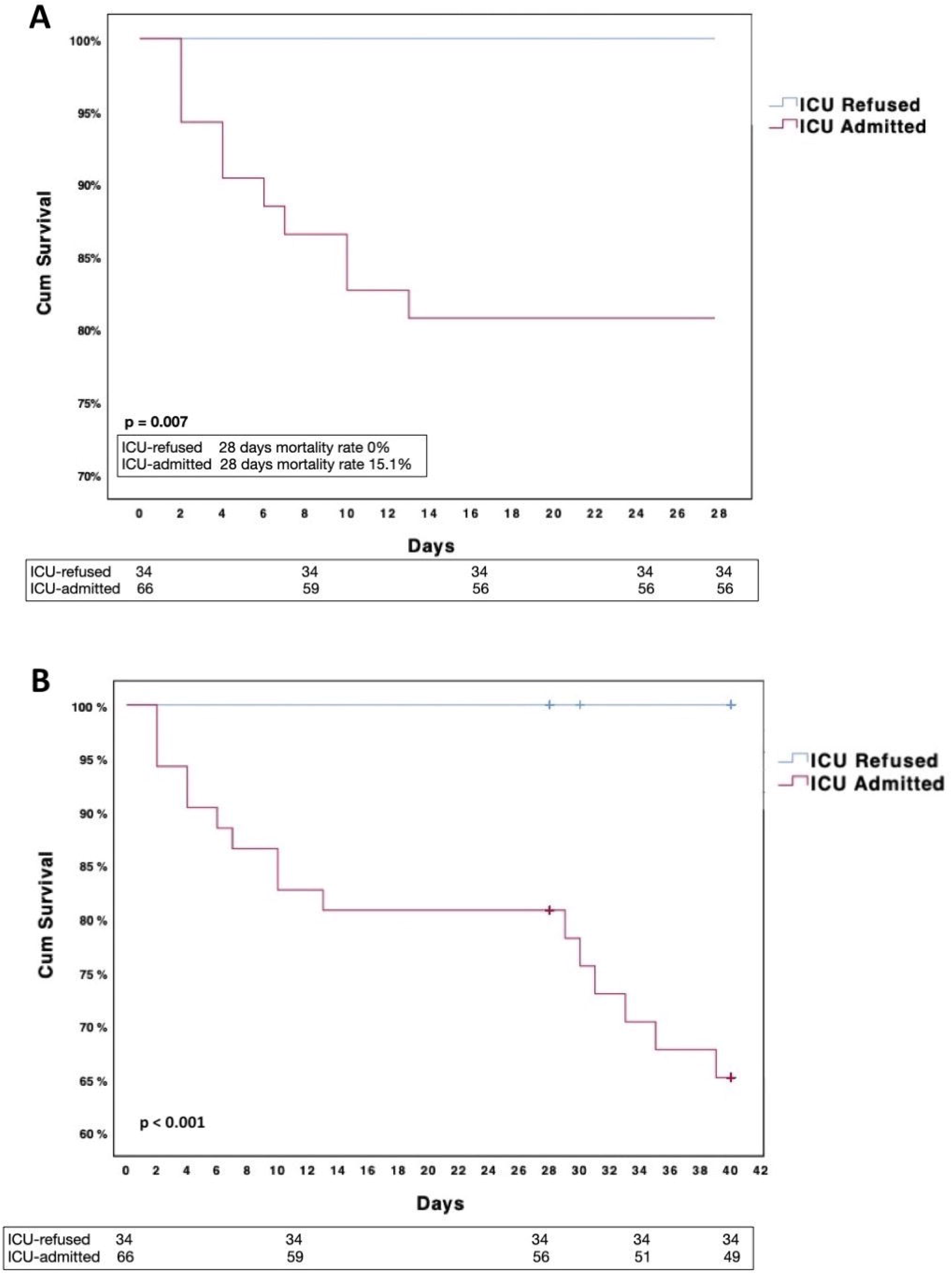
Kaplan-Meier survival at 28 days (Figure 2A) and 40 days (Figure 2B), according to outcome of Intensivist consultation (*ICU-admitted* versus *ICU-refused*), based on the presence of dyspnea and/or confusion and/or SpO_2_ less than 85%. The 28-days mortality rate resulted 15.9% in the *ICU-admitted* group and 0% in the *ICU-refused* group (Chi-square 7.195, df = 1, p = 0.007), while the 40-days mortality further increased up to 25.7% in the *ICU-admitted* group (Chi-square 13.2136, df = 1, p < 0.001).

### Hypoxia distribution rate

Using the SpO_2_ cut-off of 92%, the distribution of the hypoxia rate in both groups did not seem to correlate with ICU admission (Table 2). In the *ICU-admitted* group, 52 on 63 patients (82%) presented a SpO_2_ lower than 92% (Figure 3); similarly, 24 patients (71%) of the *ICU-refused* group presented a SpO_2_ lower than 92%. An identical distribution according to SpO_2_ cut-off value of 92% between the two groups was found (Chi-square 1.85, df = 1 p value 0.26). On the other side, using the SpO_2_ cut-off of 85% (Table 2), a correlation was found between SpO_2_ < 85% distribution and ICU-admission (Chi square 6.7, df = 1, p = 0.009). A similar correlation was also found between symptomatic dyspnea distribution and ICU-admission (Chi square 13.1, df = 1, p = 0.0002).

**FIGURE 3.**
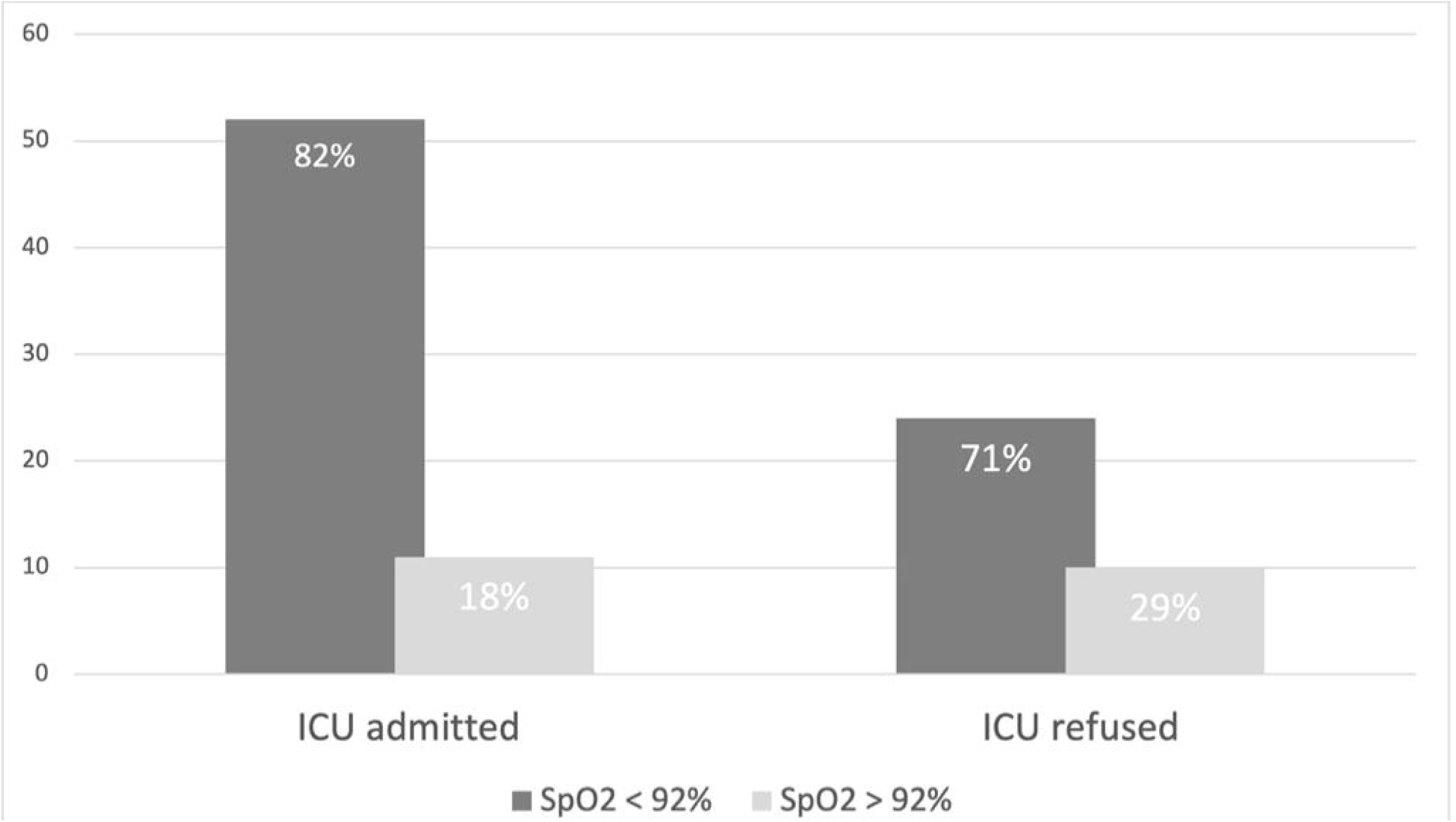
Prevalence of SpO_2_ distribution in patients at the Intensive Care consultation, stratified according to ICU admission (*ICU-admitted, ICU-refused*). In the *ICU-admitted* group, 52 of 63 patients (82%) presented a SpO_2_ less than 92%; in the *ICU-refused* group, 24 of 34 patients (71%) presented a SpO_2_ less than 92%. Chi-square analysis confirmed the identical distribution (p value = 0.26). Nearly a quarter of patients in the *ICU-refused* group presented a SpO_2_ lower than 92%, without any ICU specific inclusion criteria, and their clinical conditions progressively improved.

Using a SpO_2_ of 92% as a hypothetical cut-off allowing ICU admission, the odds ratio (OR) for patients with SpO_2_ lower than 92% compared to those with SpO_2_ greater than 92% resulted 1.96 (CI 95%, 0.73 – 5.26). Conversely, using SpO_2_ of 85% as a cut-off point, the OR to be admitted to the ICU for patients with SpO_2_ lower than 85% resulted 4.31 folds higher than those with SpO_2_ higher than 85% (CI 95%, 1.33 – 13.79). According to these data, the Bayesian post-test probability predicting model indicated that the probability to be admitted to the ICU for a SpO_2_ cut-off value of 92% is equal to 69.5%; the same probability increased up to 85.3% when the SpO_2_ cut-off value was changed to 85%.

## DISCUSSION

Acute respiratory distress induced by SARS-CoV-2 is a critical condition associated with the COVID-19 pandemic (11, 12). In order to minimize the high mortality rate associated with the disease, the adequate hospital management must also require a well-structured triage and frequent patients’ clinical evaluations (9, 13). With the aim to avoid ICU overload and to ensure simultaneously adequate medical care, we defined standardized ICU admission criteria based on partial respiratory failure with SpO_2_ lower than 85% and/or dyspnea or mental confusion. To better manage patients presenting tachypnea without dyspnea, even in the case of SpO_2_ lower than 92%, a conservative approach based on strict in-ward surveillance and regular EWS measurement (5, 8, 9) was implemented, until an eventual onset of dyspnea or of SpO_2_ below 85% allowed ICU admission. In fact, patients affected by SARS-CoV-2 interstitial pneumonia often present tachypnea and desaturation without dyspnea or neurological symptoms. An increased pulmonary compliance is probably the explanation for the absence of dyspnea in these patients (8), although further mechanisms have been proposed (14, 15). Following these criteria, none of the 34 patients not admitted to the ICU died at 28 days follow-up; all the patients improved their clinical status and all of them were subsequently discharged.

Our data suggest that in COVID-19 patients there is a low correlation between dyspnea and SpO_2_ cut-off value of 92%. The most interesting finding was that the *ICU-refused* and the *ICU-admitted* groups presented a similar SpO_2_ lower than 92% distribution. According to the patients’ distribution, the cause of ICU admission in patients with SpO_2_ greater than 92% was due to the onset of either dyspnea and/or mental confusion, confirming the lack of correlation between SpO_2_ values and the subjective feeling of shortness of breath. Analogously, there was a significant percentage of patients with SpO_2_ lower than 92% who was not admitted to the ICU, due to the absence of dyspnea and/or confusion; all of them were subsequently discharged alive and in good clinical conditions. All the collected data induce us to implement a physiology-based approach, relating to signs and/or symptoms of hypoxia together with SpO_2_ values, as the more appropriate to define ICU admission criteria. The presence of patients without hypoxia-related signs or symptoms despite SpO_2_ less than 92%, and the absence of mortality in this group, suggests that the SpO_2_ cut-off of 92% as a threshold for ICU admission in the COVID-19 context requires future re-evaluation.

A further effect of this management was the reduction of ICU-workload for healthcare professionals, which has been shown to pose a great risk for ICU-healthcare burnout (16, 17), a problem gaining uttermost importance in dramatic situations such as the COVID-19 pandemic, which are subject to lack of human and material resources. A more careful management regarding ICU-admission could allow a better management both of patients that will benefit from an intensive medical intervention than patients that do not require ICU hospitalization.

Our study was burdened by limitations. First, it was a monocentric, observational, retrospective study, with a relatively small series of patients and a lack of direct comparison with a control group. However, a stronger evaluation of our method was supplied by its application to the two different waves of COVID-19 pandemic. Second, the two groups were not completely homogeneous, differing in male sex incidence, arterial hypertension rate and serum CRP values; however, none of these values was used as criteria for decision making during the Intensivist consultation. The values may possibly be interpreted as risk factors (18) for ICU admission, rather than predictor factors; moreover, the key-message concerning the absence of mortality in the *ICU-refused* group, whose classification criteria did not involve these non-homogeneous parameters, remains intact. Finally, we are unable to define whether the SpO_2_ cut-off of 85% was the absolute best criteria to identify patients needing ICU-admission; although our study suggests that the SpO_2_ 92% threshold was unreliable in COVID-19 patients, it was not designed to specifically identify the best SpO_2_ threshold for ICU admission.

## CONCLUSIONS

In COVID-19 patients, standardized ICU admission criteria appeared to be a safe method to reduce ICU-admission, simultaneously guaranteeing a high standard of care. In the absence of dyspnea, even in the case of SpO_2_ lower than 92%, a conservative approach based on strict surveillance with regular EWS measurement in the ward was not associated with increased mortality. All collected data induce us to consider a physiology-based approach to guide ICU admission as the more appropriate during pandemics. The absence of hypoxia-related signs and/or symptoms, despite SpO_2_ less than 92%, suggests that this SpO_2_ cut-off as the threshold for COVID-19 patients’ ICU admission needs further re-evaluation.

## Data Availability

Data raw are available under written requested sent to Corresponding Author.

---

|  |  | ICU admitted | ICU refused | p value |
| --- | --- | --- | --- | --- |
| **DEMOGRAPHIC DATA** |  |  |  |  |
| Number | n (%) | 63 | 34 |  |
| Age | years | 69 ± 9 (38-89) | 70 ± 13 (31-93) | 0.87 |
| Male | n (%) | 53 (80.3) | 18 (53) | < 0.001* |
| BMI | kg/m<sup>2</sup> | 28 (24.6-32.3) | 27.9 (24.3-31) | 0.63 |
| SAPS |  | 42 (33-56) | NA | - |
| NEMS |  | 31 ± 9.5 (18-39) | NA | - |
| **COMORBIDITIES** |  |  |  |  |
| Arterial Hypertension | n (%) | 42 (66.7) | 16 (47.1) | < 0.001* |
| Ischemic cardiopathy | n (%) | 18 (28.5) | 8 (23.5) | 0.57 |
| Diabetes | n (%) | 23 (36.5) | 11 (32.4) | 0.69 |
| OSAS | n (%) | 10 (15.9) | 0 | - |
| COPD | n (%) | 9 (14.3) | 6 (17.6) | 0.59 |
| **HEMODYNAMICS** |  |  |  |  |
| Systolic arterial pressure | mmHg | 127 (115-140) | 124 (109-151) | 0.61 |
| Diastolic arterial pressure | mmHg | 65 (60-72) | 68 (57-74) | 0.58 |
| Heart Rate | bpm | 87 (77-100) | 78 (69-86) | 0.13 |
| Temperature | °C | 36.8 (36.2-37.9) | 36.4 (36-37.3) | 0.15 |
| Lactate | mmol/L | 1.6 ± 1.1 (0.5 – 6.9) | 1.2 (0.9-1.5) | 0.20 |
| **RESPIRATORY** |  |  |  |  |
| SpO<sub>2</sub> | % | 88 (55-100) | 90 (88-93) | 0.001* |
| pO<sub>2</sub> | mmHg | 57.6 (49-81) | 63.9 (56-76) | 0.07 |
| pCO<sub>2</sub> | mmHg | 35 (32-44) | 34.5 (31-39) | 0.81 |
| Hemoglobin | g/L | 13.9 ± 1.7 (9.8-17.5) | 13.4 (11.9-14.8) | 0.69 |
| O<sub>2</sub>-therapy | L/min | 15 (9.75-15) | 10 (6-13.5) | 0.02* |
| **LABORATORY** |  |  |  |  |
| ASAT | U/L | 49 (43.5-85) | 45 (34-68) | 0.97 |
| ALAT | U/L | 38 (32.5-53.5) | 35 (22-52) | 0.59 |
| Leucocyte | G/L | 9.1 ± 2 (2.3-12.3) | 5.8 (4.0-8.7) | 0.04* |
| Lymphocyte | G/L | 0.6 (0.4-0.6) | 0.7 (0.4-1.1) | 0.19 |
| C-Reactive-Protein | mg/L | 135 (116-237) | 90 (53-170) | 0.82 |
| Ferritin | ng/mL | 1781(1308-4320) | 1397 (699-2082) | 0.23 |
| LDH | U/L | 598 ± 213 (416-1048) | 375 (321-468) | 0.05* |
| Creatinine | μmol/L | 110 ± 8.9 (50-410) | 73 (60-102) | 0.23 |
| Creatinine Kinase | U/L | 266.5 (172-573) | 214 ± 330 (12-1611) | 0.26 |
| Platelets | G/L | 198 (150-254) | 264 (175-330) | 0.03* |
| Total Bilirubin | μmol/L | 9.1 (7.1-16.5) | 7.4 (5.5-10.2) | 0.21 |

Data comparison between *ICU-admitted* and *ICU-refused* groups regarding clinical and biological data. Continuous measurements are presented as mean ± SD (min-max), otherwise as median (25^th^-75^th^

|  |  |  |  |  |
| --- | --- | --- | --- | --- |
| **Dyspnea** | Yes | 41 (42.2%) | 9 (9.3%) | p = 0.0002 |
|  | No | 22 (22.7%) | 25 (25.8%) |  |
| **SpO<sub>2</sub> < 92%** | Yes | 52 (53.7%) | 24 (24.7%) | p = 0.26 |
|  | No | 11 (11.3%) | 10 (10.3%) |  |
| **SpO<sub>2</sub> < 85%** | Yes | 23 (23.7%) | 4 (4.1%) | p = 0.009 |
|  | No | 40 (41.2%) | 30 (30.9%) |  |

ICU admission criteria distribution in patients affected by COVID-19 pneumonia at the time of Intensivist consultation, according to outcome (*ICU-admitted/refused*). Regarding dyspnea, the Chi square was 13.1 (df = 1, p value 0.0002) and OR to be admitted to the ICU in case of dyspnea resulted 4.86 (CI 95%, 1.95 – 12.10). For SpO_2_ lower than 92%, the Chi-square resulted 1.85 (df = 1, p value 0.26) and the OR to be admitted to the ICU resulted 1.96 (CI 95%, 0.73 – 5.26). For SpO_2_ lower than 85%, the Chi square was 6.7 (df = 1, p value 0.009) and the OR to be admitted to the ICU resulted 4.31 (CI 95%, 1.33 – 13.79).

