## Supplementary Table 1 for "Admission criteria in critically ill COVID-19 patients: a physiology-based approach"

**SM1 Baseline characteristics between all ICU patients and patients admitted in ICU from Intensivist consultation**

|  |  | **Total ICU** |  | **ICU admitted** | **p value** |
| --- | --- | --- | --- | --- | --- |
| **DEMOGRAPHIC DATA** | | | | |  |
| Number | n (%) | 81 |  | 63 |  |
| Age | years | 68 ± 11 (29–89) |  | 69 ± 9 (38-89) | 0.67 |
| Male | n (%) | 65 (79.3) |  | 51 (80.9) | 0.87 |
| BMI | kg/m^2^ | 28 (25.2-32) |  | 28 (24.6-32.3) | 0.60 |
| SAPS |  | 47.3 ± 16.9 (13-94) |  | 42 (33-56) | 0.13 |
| NEMS |  | 34.4 ± 8.6 (9-49) |  | 31 ± 9.5 (18-39) | 0.004* |
| **COMORBIDITIES** | | | | |  |
| Arterial Hypertension | n (%) | 48 (58.5) |  | 42 (66.7) | 0.22 |
| Ischemic cardiopathy | n (%) | 20 (24.4) |  | 18 (28.5) | 0.54 |
| Diabetes | n (%) | 30 (36.6) |  | 23 (36.5) | 0.97 |
| OSAS | n (%) | 10 (12.2) |  | 10 (15.9) | 0.60 |
| COPD | n (%) | 12 (14.6) |  | 9 (14.3) | 0.86 |
| **HEMODYNAMICS** | | | | |  |
| Systolic arterial pressure | mmHg | 129 (120–140) |  | 127 (115-140) | 0.47 |
| Diastolic arterial pressure | mmHg | 65 (60–75) |  | 65 (60-72) | 0.93 |
| Heart Rate | bpm | 85 (76–96) |  | 87 (77-100) | 0.57 |
| Temperature | °C | 37.0 ± 0.9 (35.8-39) |  | 36.8 (36.2-37.9) | 0.33 |
| Lactate | mmol/L | 1.2 (0.8–1.6) |  | 1.6 ± 1.1 (0.5 – 6.9) | 0.31 |
| **RESPIRATORY** | | | | |  |
| SpO_2_ | % | 92 (88-96) |  | 88 (55-100) | 0.31 |
| paO_2_ | mmHg | 65 (52.3-87.1) |  | 57.6 (49-81) | 0.15 |
| paCO_2_ | mmHg | 35.8 (32.2-41.5) |  | 35 (32-44) | 0.34 |
| **LABORATORY** | | | | |  |
| ASAT | U/L | 47 (36-85) |  | 49 (43.5-85) | 0.72 |
| ALAT | U/L | 33 (21–48.7) |  | 38 (32.5-53.5) | 0.38 |
| Leucocyte | G/L | 7.8 ± 4.7 (2-35) |  | 9.1 ± 2 (2.3-12.3) | 0.12 |
| Lymphocyte | G/L | 0.7 (0.5–1) |  | 0.6 (0.4-0.6) | 0.23 |
| C-Reactive-Protein | mg/L | 189 ± 110 (6-534) |  | 135 (116-237) | 0.006* |
| Ferritin | ng/mL | 2379 (864–3234) |  | 1781(1308-4320) | 0.85 |
| LDH | U/L | 582 (400–720) |  | 598 ± 213 (416-1048) | 0.80 |
| Creatinine | μmol/L | 317 ± 96 (10–574) |  | 110 ± 8.9 (50-410) | 0.52 |
| Creatinine Kinase | U/L | 235 (101–367) |  | 266.5 (172-573) | 0.19 |
| Platelets | G/L | 197 ± 86 (82-458) |  | 198 (150-254) | 0.05 |
| Bilirubin total | μmol/L | 8.5 (6.5–11.9) |  | 9.1 (7.1-16.5) | 0.67 |

Data comparison between the *whole-ICU* group and the *ICU-admitted* group*,* concerning clinical and biological data. Continuous measurements are presented as mean ± SD (min-max) otherwise as median (25^th^-75^th^ interquartile) if they are not normally distributed. Categorical variables are reported as counts and percentages.

In ICU, 15 additional patients were directly transferred from other hospitals or emergency department (ED) already on MV; the group of the *whole-ICU* patients resulted composed by 81 patients. A comparison between the *ICU-admitted* and *whole-ICU* group was performed and reported in Table SM1 (supplementary material). Evaluating the *whole-ICU* group, at 28 days 43 patients (53.1%) were discharged from the ICU, eight (9.8%) still necessitated MV (6 with endotracheal tube, 2 with tracheostomy) and five (6.2%) were transferred to another hospital. At 28 days, the mortality rate of the *whole ICU* group was 30.8% (twenty-five patients).
